# Intravenous Followed by Oral Varespladib as Late Adjunctive Therapy for Snakebite: a Phase II Randomized Clinical Trial

**DOI:** 10.64898/2026.06.26.26356695

**Authors:** Charles. J. Gerardo, Ashish Bhalla, Chitta R. Mohanty, Badal K. Sahu, Surendra Kumar, Suneil Agrawal, Manu Ayyan S., Eileen Shu, Travis Deuson, Madhu Kumar R., Norman L. Beatty, Peter D. Akpunonu, Farshad M. Shirazi, Sarah Watkins, Anne-Michelle Ruha, Shawn M. Varney, Anil K. Goel, Medhavi Gautham, Justin Arnold, Chanaveerappa Bammigatti, Samuel J. Francis, Suresh Selvam, Harish Kumar, Adiel Aizenberg, Andrew Micciche, Rylee S. Bledsoe, Ginger Boatright, Suraj C. Oomman, Stephen P. Samuel, Janet T. Wittes, Rebecca W. Carter, Matthew R. Lewin, Timothy F. Platts-Mills

## Abstract

**Background:** Snakebite envenoming causes an estimated 138,000 deaths and more than 400,000 cases of permanent disability each year, disproportionately affecting low-resource regions. Antivenom is the only treatment but requires intravenous administration. We evaluated intravenous followed by oral varespladib as adjunctive therapy to antivenom in hospitalized patients with snakebite envenoming.

**Methods and findings:** In this multicenter, randomized, double-blind, placebo-controlled phase II trial in India (CTRI/2023/10/058782) and the USA (NCT05717062), patients with snakebite envenoming were randomly assigned (1:1) to receive varespladib or placebo in addition to standard of care. For elapid envenoming, the primary endpoint was time to recovery of 5-second head-lift. For viper envenoming, the primary endpoint was area under the curve (AUC) from baseline to Day 14 of a 3-item Snakebite Severity Score (SSS). Between June 3^rd^, 2023 and October 19^th^, 2024, 140 patients were randomized and 139 were analyzed (73 varespladib; 66 placebo). Study drug was initiated a mean of 7.3 hours after the bite and 3.3 hours after antivenom. Among elapid-bite patients, mean time to head-lift recovery was 27.6 hours with varespladib vs. 36.2 hours with placebo (p = 0.6). Among viper-bite patients, SSS AUC to Day 14 was 660 vs. 629 in varespladib vs. placebo (p = 0.7). No serious adverse event occurred with varespladib.

**Conclusions:** In this trial in which varespladib was initiated on average 7 hours after snakebite and several hours after antivenom, adjunctive varespladib did not improve the primary endpoints. Other study designs and settings will be needed to assess the effect of varespladib when initiated prior to antivenom.

**Synopsis:** Snakebite envenoming causes more than 100,000 deaths and hundreds of thousands of cases of long-term disability each year, primarily in low- and middle-income countries. Antivenom is the only specific treatment currently available, but it must be administered intravenously in a healthcare facility and may not fully neutralize venom toxins that have already entered tissues. Varespladib is a small-molecule drug that inhibits secretory phospholipase A2, a toxin family found in many medically important snakes. Animal studies suggest that varespladib may reduce the harmful effects of snake venom, but its effectiveness in patients remains uncertain. We conducted a randomized, placebo-controlled clinical trial in India and the United States to evaluate intravenous followed by oral varespladib as a late adjunctive treatment given in addition to standard care, including antivenom. Outcomes were analyzed in 139 patients with snakebite envenoming. Study drug began, on average, more than 7 hours after the bite and in all cases after treatment with antivenom. Varespladib was generally safe and well tolerated but did not improve the trial’s prespecified primary outcomes. These findings suggest that adjunctive varespladib does not provide an overall benefit when initiated several hours after snakebite and after antivenom administration. The study provides information to guide future research on direct toxin inhibitors for snakebite.

## INTRODUCTION

Snakebite envenoming results in approximately 150,000 deaths and 400,000 cases of permanent disability globally each year.[1] Over 50,000 of these deaths occur in India,[2] and death and disability due to snakebite envenoming are common elsewhere in the Indo-Pacific, the Middle East, Africa, and Central and South America. In the United States (U.S.), more than 10,000 people are bitten by venomous snakes annually, resulting in both short- and long-term disabilities.[3]

Antibody-based therapies (“antivenoms”) remain the only specific pharmacologic treatment currently available for snakebite envenoming. Due to the need for intravenous administration and the risk of acute allergic reactions, antivenom use is generally limited to hospital settings. Because of the rapid action of venom toxins, delays in antivenom administration are a major contributor to poor outcomes.[4, 5] An additional limitation of antivenoms is their poor tissue penetration, which results from their large size and low blood-to-tissue biodistribution coefficient.[6] This characteristic of antivenoms may reduce efficacy for pathologies such as venom-induced cytotoxicity and neuromuscular blockade.[7, 8]

Snake venom secretory phospholipase A2s (sPLA2s) are the most functionally diverse of the three most common snake venom toxins, which also include 3-finger toxins and metalloproteases.[9] Although relative abundance varies among and within species based on geography, ontogeny, and diet, [10] sPLA2s are present in the venom of at least 95% of venomous snakes worldwide, including all medically important snakes native to India and the U.S. sPLA2s act through (1) enzymatic hydrolysis of phospholipids, disrupting cellular function and releasing inflammatory mediators, and (2) in some variants, a non-enzymatic pore-forming mechanism that damages cell membranes leading to tissue damage.[11] Organ-specific injury occurs via sPLA2 binding to proteins on target cells.[12] Because of their small size, which allows for rapid diffusion and systemic effects, and their effects on neuromuscular and neurovascular function, sPLA2s contribute to lethal effects of snakebite envenoming; hence, venoms with a higher sPLA2 content tend to be more toxic.[10] The prevalence, functional diversity, and role in severe pathologies of snakebite envenoming make this toxin family a promising therapeutic target.[13, 14]

Varespladib, a potent small molecule inhibitor of snake venom sPLA2, can be administered intravenously as a sodium salt (LY351920) and orally as varespladib methyl (LY333013, *syn* S3013). In preclinical studies in mice and pigs, varespladib administered shortly after envenoming results in either delayed lethality or increased survival following exposure to lethal doses of venom from the elapids and vipers included in this study: (1) krait (*Bungarus spp.*) and eastern coral snake (*Micrurus fulvius*);[15, 16] (2) U.S. Crotalinae including copperhead (*Agkistrodon contortrix*), western diamondback (*Crotalus atrox*), and pygmy rattlesnake (*Sistrurus miliarius*), as reported in a recent pre-print publication;[17] and (3) Russell’s viper (*Daboia russelii*), though efficacy varies regionally across India due to variations in the venom proteome of this species.[18] A prior clinical trial of varespladib for snakebite envenoming, the BRAVO trial, used a similar hospital-based design with patients receiving standard of care including antivenom but tested an oral-only dose regimen.[19] Results suggested improved outcomes from varespladib vs. placebo in patients who received the study drug within 5 hours of snakebite.

Although early field or prehospital varespladib has been recognized as the use case with the greatest potential public health benefit, such trials are logistically challenging. Thus, to advance understanding of varespladib for snakebite, this trial assessed the efficacy, safety, and tolerability of intravenous varespladib sodium followed by oral varespladib methyl when given in addition to standard of care (including antivenom) in hospitalized patients with snakebite envenoming.

## METHODS

### Trial Design and Participants

We conducted this multicenter randomized, double-blind, placebo-controlled trial at 8 sites in India and 10 in the U.S. (Appendix 1). Sites were selected to provide broad geographic representation and capture a diverse range of medically important snake species and envenoming syndromes across both countries. Site selection also prioritized investigators with established expertise in snakebite management and clinical research. The trial protocol was approved by Indian and U.S. regulatory authorities and the Institutional Review Boards (IRB) or Ethics Committees (EC) at each site. An independent data and safety monitoring board oversaw and periodically reviewed safety data from the trial. All sites received the study protocol and pharmacy manual and underwent a site initiation visit before enrollment began. The contract research organization overseeing the trial monitored sites to review procedures and ensure protocol compliance and data quality. None of the funding agencies controlled the trial design or conduct, analysis of the data, or writing of the manuscript. The trial was registered at ClinicalTrials.gov, NCT05717062 (https://clinicaltrials.gov/study/NCT05717062), and Clinical Trials Registry-India, CTRI/2023/10/058782 (https://ctri.nic.in/Clinicaltrials/pmaindet2.php?EncHid=ODI4NTI=&Enc=&userName=).

All patients received standard of care, including antivenom as indicated clinically and consistent with national guidelines in India and manufacturer recommendations in the U.S., without any delays or restrictions. Patients 18 years of age or older who presented to emergency departments with signs and symptoms of snakebite envenoming were screened for eligibility. In the U.S., the age range was extended to age 5 years and older on 13 August 2024. Pediatric enrollment was introduced later in the trial because additional regulatory review was required to establish an appropriate pediatric dosing regimen for varespladib. By the time pediatric enrollment was approved in the U.S., enrollment in the trial was nearing completion and pediatric enrollment was not pursued in India.

In the U.S., there was no exclusion based on the species of the offending snake or inability to identify the snake. In India, patients were eligible if they had suspected or confirmed bites from krait (*Bungarus spp.*) or Russell’s viper (*Daboia russelii*). Because snake species is a major determinant of venom composition, clinical manifestations, severity, and outcomes of envenoming, enrollment in India was restricted to these species to reduce clinical and venom-related heterogeneity and facilitate interpretation of treatment effects. Russell’s viper and krait were selected because they are common medically important causes of envenoming in India and were the most frequently enrolled snake types in the preceding BRAVO trial. In both the U.S. and India, patients with viper-bites were eligible if they were randomized within 5 hours of bite or symptom onset and had a Snakebite Severity Score (SSS) of 3 or more based on the composite of the local wound, pulmonary, cardiovascular, hematologic, and nervous system subscores (Appendix 2).[20] Patients with elapid bites were eligible if randomized within 10 hours of bite or symptom onset and had moderate or severe cranial nerve or skeletal muscle weakness. Site investigators made the diagnosis of snakebite envenoming. Snake species were determined based on photographs when available, clinical assessment, and local species epidemiology. Patients were excluded if they had a history of cerebrovascular accident, acute coronary syndrome, chronic kidney disease, or chronic liver disease. See Appendix 3 for a full list of eligibility criteria.

Changes to the protocol were implemented following external review of results from a separate, related study (the BRAVO trial). These included: (1) increasing the sample size from 110 to 140 patients; (2) restricting inclusion criteria in India to patients bitten by Russell’s viper and krait; and (3) revising the primary endpoint from change in Snakebite Severity Score (SSS) from baseline to the average at 3 and 6 hours to separate endpoints for elapid and viper envenomation, as described below. These modifications were prospectively specified in a protocol amendment prior to database lock and were made while the study remained blinded, with no access to unblinded BRAVIO data.

### Randomization and Allocation Concealment

Patients were block randomized in a 1:1 ratio to varespladib or placebo with concealed allocation. Randomization was stratified by snake type (copperhead, Mojave rattlesnake, non-Mojave rattlesnake, Russell’s viper, Elapid, or other) and by age group (ages 5-17 or ≥18 years). A computer-generated list was used for randomization. The treating team, trial management group, and patients remained unaware of group assignments until the final analysis of the study was complete. Intravenous study drug was provided to the clinical team by an unblinded research pharmacist in the form of a 250 mL bag of normal saline with or without varespladib sodium. The bag and intravenous line had an opaque cover to prevent potential unblinding resulting from the slight coloration of varespladib sodium. Research pharmacists who prepared the intravenous study drug were not otherwise involved with the study.

### Intervention

All patients received standard of care, including antivenom without any restrictions on time to initiation. Adult patients randomized to the active drug received a constant rate infusion of 0.45 mg/kg/hr of varespladib sodium for 6 hours. At 6 hours, patients who were clinically stable and able to tolerate oral medication were switched to varespladib methyl with a 500 mg loading dose followed by 250 mg twice a day for a total treatment duration of 7 days. Patients who were unable to take oral medication at 6 hours were continued on the intravenous infusion until they were able to switch. The dosing for pediatric patients was 0.6 mg/kg/hour intravenous infusion followed by an age-based dose of the oral drug following the dosing in the BRAVO trial.[19] Patients randomized to placebo received an intravenous infusion of normal saline and the equivalent number of oral placebo pills. Both varespladib methyl and placebo tablets and capsules were made by the same manufacturer under Good Manufacturing Practices conditions.

### Outcomes

For elapid-bite patients, the primary outcome was the elapsed time from randomization to recovery of 5-second head-lift, defined by a head-lift of 5 seconds. This outcome was selected because it was familiar to clinicians, clinically relevant, and expected to improve with reversal of venom-induced paralysis. For viper-bite patients, the primary outcome was the area under the curve from baseline to Day 14 for a 3-item SSS composed of the local wound, hematologic, and neurologic subscores. This outcome was selected because it captures the cumulative burden of illness over time across the major clinically relevant manifestations of viper envenoming and is sensitive to differences in both the magnitude and duration of toxicity. Day 14 was selected because clinically important manifestations of viper envenoming, particularly local tissue injury, often persist beyond the acute phase and may continue to evolve after correction of systemic toxicity and coagulopathy. The global primary endpoint was to be tested by combining the p-values from these two outcomes using Fisher’s combined probability test.

Three alpha-protected key secondary outcomes were identified: (1) area under the curve (AUC) for the 6-component SSS (local wound, pulmonary, cardiovascular, hematologic, nervous, and renal system subscores) from baseline to Day 7 among the subgroup of patients randomized within 5 hours of the bite; (2) Complete recovery of the SSS at Day 28, defined by a score of zero for the 6-component SSS; and (3) Patient Specific Functional Scale (PSFS) at Day 3 (48 hours). The Holm-Bonferroni method was used to control the Type I error rate.

Safety was assessed with vital signs, physical examination, laboratory tests, electrocardiograms (EKGs), clinician assessments, and patient-reported symptoms and categorized by system organ class and preferred term using the Medical Dictionary for Regulatory Activities. Concomitant medication and therapies, interventions, duration of hospitalization, and adverse events were also recorded. Patient input informed the selection and timing of outcome assessments in the trial.

### Sample Size

We determined that the enrollment of 40 patients bitten by elapids and 100 patients bitten by vipers would provide the study with over 90% power to detect a statistically significant difference between varespladib and placebo, based on a global combined p-value with a two-sided Type 1 error rate of 5%. The sample size for the elapid-bite patients, which assumed a distribution of times to head-lift recovery based on the BRAVO data (Appendix 4), was calculated using a randomization test with 200 simulations. The sample size for viper-bite patients was based on a clinically meaningful difference of 125 points, a standard deviation of 225, and a requirement of 80% statistical power.

### Statistical Analysis

The primary analysis was performed in the intention-to-treat (ITT) population. Missing data were imputed (Appendix 5). The ITT included all randomized patients except for one patient who was erroneously given placebo for the infusion treatment phase instead of active drug. The patient then received active oral drug as assigned. Safety data are reported separately for this patient.

For elapid-bite patients, time to complete head-lift recovery was compared between treatment groups using a randomization test with baseline head-lift as a covariate in two categories: 0 seconds and 1 to 4 seconds. The randomization test was selected because of the likelihood of very unequal variances between the treatment groups. This analysis excluded patients with a head-lift of 5 seconds (i.e., normal or fully recovered) at baseline. The three key secondary efficacy endpoints—AUC of the 6-component SSS through Day 7 (≤5-hour subgroup), complete SSS recovery at Day 28, and PSFS score at Day 3—were analyzed in the intention-to-treat population using prespecified models aligned with the statistical analysis plan. Continuous endpoints (SSS AUC and PSFS) were compared between treatment groups using analysis of covariance (ANCOVA) adjusting for baseline severity and stratification factors, while complete SSS recovery was analyzed using a logistic regression model with similar covariate adjustment. Missing data were handled using multiple imputation under a treatment-policy estimand, and multiplicity across key secondary endpoints was controlled using a hierarchical testing procedure (Holm-Bonferroni). For viper-bite patients, the SSS AUC Day 14 was compared between treatment groups using analysis of covariance with snake type (copperhead/water moccasin, rattlesnake, or Russell’s viper) as a covariate. For this covariate, the patient bitten by a water moccasin was included with the copperhead-bite patients. However, consistent with the statistical analysis plan, subgroup analyses of patients bitten by copperheads do not include the patient bitten by a water moccasin. Pre-planned subgroup analyses by country, snake type, and time from bite to randomization are applied to the key secondary outcomes, including prespecified subgroup analysis for patients bitten by copperheads. Adverse events were compared between treatment groups. Post-hoc analyses were conducted to further characterize the effect of varespladib vs. placebo on outcomes in the subgroup of patients bitten by specific snake types.

All analyses were performed with SAS version 9.4.

## RESULTS

### Recruitment and Participant Flow

From June 3^rd^, 2023 through October 19^th^, 2024, a total of 557 patients were assessed for eligibility. The most common reasons for exclusion were failure to meet the severity criteria or not meeting the eligibility window (Figure). The intent-to-treat population of 139 patients was analyzed for efficacy outcomes. Outcome assessments were completed for 135 (97%) patients on Day 2, 134 (96%) patients on Day 7, and 135 (97%) patients on Day 28.

**Figure:**
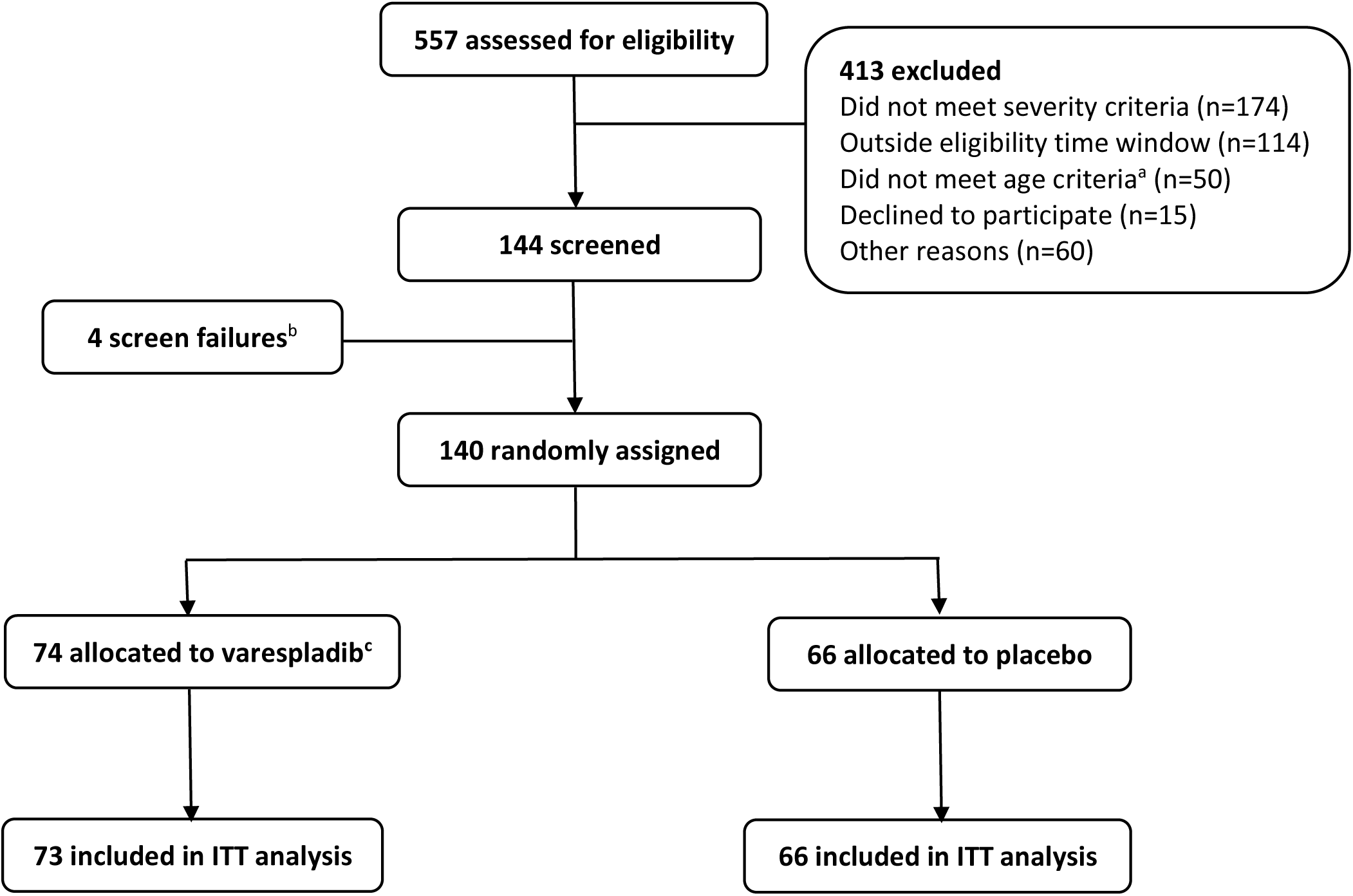
Trial enrollment ITT = intention-to-treat ^a^ Prior to 31 July 2024 patients age 18 and older were not eligible in India and the U.S.; after 31 July 2024 pediatric patients aged 5 to 17 years were also eligible for U.S. sites. ^b^ Screen failures: Patient gave verbal consent but then declined to sign consent (n=1); Patient withdrew consent before dosing (n=1); Patient did not meet eligibility criteria (n=2). ^c^ One patient randomized to varespladib, but received placebo IV treatment, then varespladib oral treatment. This patient was excluded from the primary analysis.

### Baseline Data

Baseline characteristics were well balanced between treatment groups (Table 1). Eighty-six patients were enrolled in India; 53 patients were enrolled in the U.S. For the entire sample, the mean time from bite to initiation of antivenom was 4 hours and 15 minutes and from bite to initiation of study drug was 7 hours and 18 minutes, a delay of 3 hours 3 minutes from antivenom administration. Among patients bitten by vipers, the mean time from bite to initiation of study drug was 6 hours and 8 minutes.

**Table 1.** Baseline characteristics.

| Characteristic | Varespladib<br>(n=73) | Placebo<br>(n=66) |
| --- | --- | --- |
| Age |  |  |
| Mean (SD) – yr | 41 (16) | 43 (16) |
| 5-17 years – no. (%) | 1 (1) | 0 |
| 18 years and older – no. (%) | 72 (99) | 66 (100) |
| Sex |  |  |
| Male – no. (%) | 52 (71) | 48 (73) |
| Female – no. (%) | 21 (29) | 18 (27) |
| Country |  |  |
| India – no. (%) | 44 (60) | 42 (64) |
| United States – no. (%) | 29 (40) | 24 (36) |
| Time from bite in hours |  |  |
| Bite to antivenom – mean (SD) | 4.3 (3.5) | 4.0 (3.1) |
| Bite to study drug – mean (SD) | 7.5 (3.7) | 7.1 (3.5) |

| Snake type – no. (%) |  |  |
| --- | --- | --- |
| Krait | 24 (33) | 21 (32) |
| Eastern coral snake | 0 | 1 (2) |
| Russell's viper | 20 (27) | 21 (32) |
| Rattlesnake<sup>a</sup> | 16 (22) | 11 (17) |
| Copperhead | 12 (16) | 12 (18) |
| Water moccasin | 1 (1) | 0 |
| Baseline Snakebite Severity Score |  |  |
| Mean (SD) | 6 (2) | 6 (2) |
| Range | 2-14 | 2-11 |

<sup>a</sup> *Crotalus horridus*, *C. atrox*, *C. helleri*, *C. cerastes*, *C. scutulatus*, *Sistrurus miliarius*, and unknown.

Although patients bitten by vipers had to be enrolled within 5 hours of bite or symptom onset, the time from bite to initiation of study drug was often more than 5 hours because of the time needed to obtain the study drug from the pharmacy. Additionally, some patients reported symptoms onset that occurred several hours after the bite; these patients had a time from symptom onset to enrollment within 5 hours but a time from bite to enrollment that was more than 5 hours.

Among the 137 patients in the ITT population who received antivenom, 132 patients (96%) received antivenom prior to receiving study drug. The mean time from antivenom initiation to study drug initiation was 3 hours and 17 minutes. In the U.S, patients received crotalidae polyvalent immune Fab (ovine) (CroFab) and/or crotalidae immune F(ab’)₂ (equine) (Anavip) fractionated antivenom. In India, patients received VINS, Premium Serums, or Bharat Serums antivenoms, each of which is an F(ab’)2. In 82% of cases, the baseline SSS was assessed after the patient had received an initial dose of antivenom. Two patients, both in the U.S., did not receive antivenom: One was enrolled at University of Florida, Gainesville, FL and bitten by a pygmy rattlesnake (*Sistrurus miliarius*) and one was enrolled at the University of Texas Health-San Antonio, San Antonio, TX and bitten by an unknown species of rattlesnake. Both patients received varespladib.

### Primary and Key Secondary Outcomes

For elapid-bite patients, the adjusted mean (SE) time to head-lift recovery was 27.6 (10.5) hours in varespladib-treated patients vs. 36.2 (11.1) hours in placebo-treated patients (p = 0.6). For viper-bite patients, the adjusted mean SSS AUC Day 14 was 660 (54.3) in varespladib-treated patients vs. 629 (57.1) in placebo-treated patients (p = 0.7). Consequently, the global primary endpoint was not calculated. The three key secondary outcomes were also not statistically different by treatment group (Table 2).

**Table 2.** Primary and key secondary outcomes for the intention-to-treat population (n=139).

| Outcome | Varespladib<br>(n=73) | Placebo<br>(n=66) | Treatment Effect<br>(95% CI) |
| --- | --- | --- | --- |
| Primary outcome, mean (SE) |  |  |  |
| Elapids – Time to complete head-lift recovery (hrs)<sup>a</sup> | 27.6 (10.5) | 36.2 (11.1) | -8.6 (-38.3 to 21.1)<br>(favors varespladib) |
| Vipers – AUC Day 14 of 3-item SSS<sup>b</sup> | 660 (54) | 629 (57) | 30 (-123 to 183)<br>(favors placebo) |
| Key secondary outcomes |  |  |  |
| AUC Day 7 of 6-item SSS for patients randomized $\leq 5$ hrs from bite, mean (SE) | 467 (39) | 465 (44) | 2 (-103 to 107)<br>(favors placebo) |
| Complete SSS Recovery at Day 28 | 55.4% | 51.5% | 3.9% (-12.7% to 20.5%)<br>(favors varespladib) |
| PSFS score on Day 3, mean (SE) | 6.5 (0.4) | 6.1 (0.4) | 0.4 (-0.6 to 1.4)<br>(favors varespladib) |

AUC = area under the curve; SSS = Snakebite Severity Score; SE = standard error; PSFS = patient-specific functional scale
<sup>a</sup> Varespladib n = 22 (with baseline 5-second head-lift $< 5$ ); Placebo n = 20 (with baseline 5-second head-lift $< 5$ ).
<sup>b</sup> Varespladib n = 49; Placebo n = 44

### Results for Elapids

For patients bitten by elapids, several additional outcomes indicate clinically important differences favoring varespladib (Table 3). Illness severity over the first week as measured by the SSS AUC Day 7 was 36% lower in varespladib-treated patients (nominal p = 0.003). Among intubated patients (n = 25), the duration of intubation and mechanical ventilation was 21 hours vs. 40 hours in varespladib- vs. placebo-treated patients (nominal p = 0.02). The improvement in the 5-item SSS from baseline to the average at 3 and 6 hours was greater in varespladib-treated patients (nominal p = 0.04)

**Table 3.** Outcomes in patients bitten by elapids (n=46).

| Outcome | Varespladib<br>(n=24) | Placebo<br>(n=22) | Treatment Effect<br>(95% CI) |
| --- | --- | --- | --- |
| Primary outcome |  |  |  |
| Time to complete head-lift recovery, mean (SE) | 27.6 (10.5) | 36.2 (11.1) | -8.6 (-38.3 to 21.1)<br>(favors varespladib) |
| Key secondary outcomes, Elapid subgroup |  |  |  |
| AUC Day 7 of 6-item SSS, mean (SE)<sup>a</sup> | 295 (38) | 458 (40) | -163 (-271 to -55)<br>(favors varespladib) |
| Complete SSS Recovery at Day 28 | 58.3% | 45.5% | 12.8% (-16.8 to 41.6%)<br>(favors varespladib) |
| Patient Specific Functional Scale Day 3, mean (SE) | 8.4 (0.6) | 7.1 (0.7) | 1.3 (-0.5 to 3.1)<br>(favors varespladib) |
| Exploratory outcome |  |  |  |
| Duration of intubation, mean (SE) | 21 (5)<sup>b</sup> | 40 (5)<sup>b</sup> | -19 (-33 to -4)<br>(favors varespladib) |
| Post-hoc outcome |  |  |  |
| Change in 5-item SSS from Baseline to the<br>averaged at 3 and 6 hours, mean (SE)<sup>c</sup> | -1.2 (0.3) | -0.4 (0.3) | -0.8 (-1.6 to -0.05)<br>(favors varespladib) |

AUC = area under the curve; SSS = Snakebite Severity Score; SE = standard error
<sup>a</sup> Among elapid-bite patients, only 3 patients were treated ≤5 hours from bite. Results are shown for all elapid-bite patients.
<sup>b</sup> Intubated varespladib patients = 12, Intubated placebo patients = 13
<sup>c</sup> Adjusted for baseline 5-item SSS.

### Results for Vipers

Among all viper-bite patients, neither the primary endpoint nor the key secondary endpoints were statistically different by treatment group (Table 4). The two patients bitten by rattlesnakes who received varespladib but did not receive antivenom both had complete recovery (SSS = 0) by Day 14.

**Table 4.** Outcomes in patients bitten by vipers (n=93).

| Outcome | Varespladib<br>(n=49) | Placebo<br>(n=44) | Treatment Effect<br>(95% CI) |
| --- | --- | --- | --- |
| Primary outcome |  |  |  |
| 3-item SSS AUC Day 14 | 660 (54) | 629 (57) | 30 (-123 to 183)<br>(favors placebo) |
| Key secondary outcomes, Viper subgroup |  |  |  |
| 6-item SSS AUC Day7 | 480 (31) | 461 (33) | 19 (-69 to 107)<br>(favors placebo) |
| Complete recovery at Day 28, n (%) | 53.9% | 54.6% | -0.7% (-21.8 to 18.8%)<br>(favors placebo) |
| Patient Specific Functional Scale Day 3, Mean (SE) | 6.3 (0.7) | 6.4 (0.8) | -0.1 (-2.3 to 2.0)<br>(favors placebo) |

### Ancillary Analyses

Outcomes for the key secondary endpoints did not differ statistically between treatment groups when examined separately for pre-specified subgroups: patients in India and U.S. or time from bite to treatment ≤5 hours and >5 hours (Appendix 6). Although 66 patients were randomized ≤5 hours from the bite, only 36 patients (19 varespladib and 17 placebo) received the study drug in ≤5 hours from the bite.

Among patients bitten by copperheads (n=24), as compared to placebo, varespladib-treated patients had lower illness severity over the first two weeks (3-item SSS AUC from baseline to Day 14; viper primary endpoint; nominal p = 0.04) and lower illness severity over the first week (6-item SSS AUC from baseline to Day 7; nominal p = 0.04), with the point estimates for the viper primary endpoint indicating a 43% reduction in illness severity with varespladib treatment. Mean pain scores at Day 3 (48 hours post enrollment) were 1.5 points lower in varespladib-treated patients (nominal p = 0.08). From baseline to the average at 3 and 6 hours, varespladib treated-patients showed a greater improvement in the 5-item SSS compared to placebo, but this trend was not statistically significant (nominal p = 0.10).

### Safety Outcomes

No death, serious adverse event, or investigator request to discontinue study drug or unblind a patient occurred in varespladib-treated patients. No case of stroke, acute coronary syndrome, or malignant cardiac arrhythmia occurred, and no patient was found to have EKG changes to suggest cardiac ischemia at any time point. There were no differences in vital signs between the treatment groups. Two serious adverse events occurred in patients receiving placebo, both in patients bitten by copperheads.

One patient had evidence of infection at the bite site on his thumb on study Day 6 that required re-hospitalization, debridement, and intravenous antibiotics. This patient ultimately required surgical repair of venom-induced injury of the extensor tendon of his first digit of his left hand. The second serious adverse event occurred in a patient with a history of opioid use disorder who experienced an opioid overdose on study Day 27. During the initial hospitalization, the patient had severe pain from the bite that required treatment with opioids.

The percentage of patients experiencing one or more treatment-emergent adverse event was 26% and 30% in the varespladib and placebo groups, respectively (Appendix 7). Possibly-related treatment emergent adverse events were reported in 4% vs. 11% of varespladib and placebo patients, respectively. Gastrointestinal disorders occurred infrequently in varespladib-treated patients, and no case of nausea or vomiting occurred. The frequency of other adverse events was similar in the varespladib- and placebo-treated patients. Elevated creatinine was observed at a similar frequency in varespladib- and placebo-treated patients (10% vs. 9%), and in all varespladib-treated patients with elevated creatinine, creatinine had returned to normal by Day 28. The percentage of patients receiving varespladib who had an elevation in liver enzymes was lower than the percentage of placebo patients (3% vs. 12%). The two varespladib-treated patients with elevated liver enzymes also had elevations in creatinine kinase, suggesting that the liver enzyme elevation was due to cytotoxicity secondary to snakebite envenoming and not a result of liver injury from the study drug. The patient who was randomized to varespladib but erroneously received IV placebo treatment, then oral varespladib, experienced cough, fever, and nasopharyngitis on Day 6. These adverse events were reported to be mild and unrelated to the study drug.

### Pharmacokinetic Results

Patients randomized to varespladib had median serum varespladib levels at 30 minutes of 1510 ng/mL and at end of infusion of 1840 ng/mL. Two hours after the loading oral dose, the median varespladib drug level was 1030 ng/mL, and at 5 to 10 hours was 668 ng/mL. Prior to dosing on Day 7, the median drug level was 231 ng/mL, indicating little or no accumulation over the 7-day dosing period.

## DISCUSSION

In this two-country, multicenter, randomized clinical trial of patients receiving in-hospital treatment for snakebite envenoming, adjunctive varespladib did not meet the primary superiority endpoint compared with placebo when added to standard of care. Several aspects of the design may have limited the ability to detect a treatment effect. Patients initiated study drug several hours after envenoming, in part because preparation of the intravenous study drug by an unblinded research pharmacist was required before administration. In addition, consistent with standard management of snakebite envenoming, patients received antivenom promptly after hospital presentation and before initiation of study drug.

Consequently, this trial evaluated adjunctive varespladib administered after antivenom and does not inform the potential efficacy of varespladib administered earlier, including as a pre-referral or prehospital therapy before antivenom is available.

Venom compositions and toxicities may also have impacted the findings from the trial. In BRAVIO, the two most common snakebites in India and the U.S. were Russell’s viper and rattlesnakes. sPLA2 is a lethal venom component in these snakes, and preclinical studies have demonstrated that varespladib improves survival and mitigates toxicity in animal models of medically important U.S. rattlesnakes and Russell’s viper when administered early after venom exposure.[17, 18] However, none of the patients in either treatment group died. In both rattlesnake and Russell’s viper envenoming, coagulopathy is common and is often effectively treated with antivenom, which may reduce the incremental benefit of sPLA2 inhibition when administered later in the clinical course.[21] In addition, both snake groups exhibit substantial geographic and species-level variation in venom proteome, including variable contributions of metalloproteases and other non-sPLA2 toxins that may limit the effectiveness of sPLA2 inhibition alone.[18, 22] Together, these considerations suggest that earlier initiation of therapy and, in some contexts, multi-target toxin inhibition strategies may be necessary to realize the full clinical potential of direct toxin inhibitors.

Pre-specified subgroup analyses suggest clinically important benefits of varespladib among patients bitten by kraits and copperheads; these findings warrant further evaluation. Several features support the biological and clinical plausibility of these observations: improvements occurred rapidly, within 6 hours of treatment initiation; effect sizes were substantial, including approximately 35% lower illness severity over the first week and a 47% shorter duration of mechanical ventilation among intubated krait-bite patients; these species have well-characterized sPLA2-mediated toxicities;[23, 24] and treatment effects were observed across multiple clinically relevant endpoints. These findings suggest that varespladib may mitigate both neurotoxicity, as reflected in the krait subgroup findings, and soft-tissue injury, as reflected in the copperhead subgroup findings, despite concurrent administration of standard-of-care antivenom. The reduction in ventilation duration among krait-bite patients may be particularly important in resource-limited settings,[25] while improvements in illness severity and pain among copperhead-bite patients could translate into reduced short-term disability and earlier return to normal activities. The observed subgroup findings in these patients who have already received antivenom may relate to the limited tissue penetration of antibody-based antivenoms compared with the small molecular size of varespladib, which may more rapidly access muscle and other tissues where sPLA2-mediated injury occurs.[6, 7] Variability in antivenom recognition of sPLA2 toxins may also contribute to incomplete neutralization in some settings.[26]

Unlike the BRAVO trial, this study did not demonstrate a clear interaction between treatment effect and time to initiation of therapy, possibly because few patients received study drug very early after envenoming. Delays in reaching hospital-based antivenom remain a major contributor to poor outcomes globally, and based on animal studies, venom kinetics, and epidemiologic studies,[27] we think that, like antivenom, time to treatment will influence the degree of benefit from varespladib. Further evaluation of varespladib in settings that enable earlier administration are needed.

The BRAVIO trial provides further evidence that varespladib is generally safe and well tolerated for the treatment of snakebite. No patient experienced a severe or serious adverse event, and even among patients with higher exposures during the IV phase, the rate of adverse events was similar to those receiving placebo.

This study has several limitations. First, because study drug was initiated a mean of more than 7 hours after snakebite and after antivenom, the results do not inform the efficacy of varespladib when administered earlier, including as a prehospital or pre-referral treatment. Second, antivenom type, dose, and timing were not standardized because treatment followed local practice. Differences in antivenom products and their neutralizing activity may have contributed to variability in clinical response and reduced the ability to detect an incremental benefit of adjunctive varespladib. Third, although several subgroup analyses were prespecified, the numbers of patients within individual snake groups were modest and the study was not powered to detect species-specific treatment effects. These findings should therefore be interpreted cautiously and require confirmation in future studies.

In this study of hospitalized patients receiving antivenom for snakebite envenoming, late adjunctive varespladib did not improve the prespecified primary endpoints in the overall population. Other study designs are needed to evaluate the effect of field or prehospital varespladib as a pre-referral treatment for snakebite.

## Author Contributions

CJG, AB, SPS, RWC, MRL and TPM were responsible for the original study concept and design. TPM, RWC, and JW developed the statistical analysis plan. TPM, SPS, GB, and SCO contributed to the coordination of the study. CJG, AB, CRM, BKS, SK, SA, MAS, ES, MKR, NLB, PDA, FMS, SW, RM, SMV, AKG, MG, JA, SJF, SS, AA, AMR, AM screened and enrolled patients into the trial, and participated in data collection, data analysis, and review of the manuscript. TPM, SCO, and SPS contributed to the clinical data review. CJG, AB, and TPM had full access to all data in the study and take responsibility for the integrity of the data and the accuracy of the data analysis and serve as guarantors for the manuscript. All authors read and approved the final manuscript.

## Ethical Approvals

This work was reviewed by FDA (IND 142870, Protocol OPX-PR-03 v1.0, dated January 28, 2022, submitted February 11, 2022, Serial/Sequence Number 0005) in U.S. and the CDSCO (CT/22/000169) in India. The study was also reviewed and approved by the WCGIRB (Tracking Number 20226769) in the U.S. and by the following Ethics Committees in India: S. P. Medical College, Bikaner (Approved on 19 Sep 23 ref EC/SPMC/ECA/094), PGIMER, Chandigarh (Approved 23 Aug 23 PGI/IEC/CT/2023/00933), JIPMER, Pondicherry (20 Mar 24 JIP/IEC/2024/03/34), K. R Hospital (Approval on 10 April 2023 MMC EC 31/23), All India Institute of Medical Sciences, Raipur (Approved 24 Apr 23 AIIMSRPR/IEC/2023/1350), All India Institute of Medical Sciences, Bhubaneshwar (10 Jan 24 T/EMF/T&EM/2023-24/193), King George’s University Hospital, Lucknow (16 Apr 24 149/Ethics/2024 or 130th ECMIIC/P2), and Nil Ratan Sircar Medical College & Hospital, Kolkata (Approved 15 Jan 24 NRSMC/IEC/08/2024) from India.

## Patient Consent

Written informed consent was obtained from all patients or legally authorized representatives for those unable to consent themselves.

## Data Availability

De-identified data and analysis code are available via Zenodo (https://doi.org/10.5281/zenodo.19075404). The dataset has been de-identified in accordance with applicable regulatory standards to protect participant privacy. Supporting documentation necessary to interpret the data, including data dictionaries and metadata, is provided.

## Acknowledgments

We thank the patients for their willingness to participate in the study, those within our organization and at study sites for the dedication required to successfully perform this study, and the DSMB members for their oversight of the study.

---

**Appendix 1.** List of sites in India and the U.S. for the BRAVIO trial.

**Appendix 2.** Snakebite Severity Score versions used for (i) Inclusion Criteria; (ii) Primary Outcome (Viper); (iii) Secondary Outcomes (SSS AUC basline to Day 7 and complete recovery).

**Appendix 3:** Complete eligibility criteria.

**Appendix 4:** Details of the sample size calculation for patients bitten by elapids.

**Appendix 5:** Multiple imputation methodology.

**Appendix 6:** Key secondary outcomes in pre-specified subgroups.

**Appendix 7.** Adverse events and selected laboratory abnormalities. Values represent the number (%) of patients with one or more adverse events or laboratory abnormalities.

## REFERENCES

1. Warrell, D.A. and D.J. Williams, Clinical aspects of snakebite envenoming and its treatment in low-resource settings, in The Lancet. 2023, Elsevier B.V. p. 1382–1398.

2. Suraweera, W., et al., Trends in snakebite deaths in India from 2000 to 2019 in a nationally representative mortality study. eLife, 2020. 9: p. 1–37.

3. Jaramillo, J.D., et al., The “T’s” of Snakebite Injury in the USA: Fact or Fiction? Trauma Surgery & Acute Care Open, 2019. 4(1): p. e000374–e000374.

4. Mohapatra, B., et al., Snakebite Mortality in India: A Nationally Representative Mortality Survey. PLoS Neglected Tropical Diseases, 2011. 5(4): p. 1–8.

5. Habib, A.G. and S.B. Abubakar, Factors Affecting Snakebite Mortality in North-Eastern Nigeria. International Health, 2011. 3(1): p. 50–55.

6. Shah, D.K. and A.M. Betts, Antibody biodistribution coefficients: inferring tissue concentrations of monoclonal antibodies based on the plasma concentrations in several preclinical species and human. MAbs, 2013. 5(2): p. 297–305.

7. Prasarnpun, S., et al., Envenoming bites by kraits: the biological basis of treatment-resistant neuromuscular paralysis. Brain, 2005. 128: p. 2987–2996.

8. Bartlett, K.E., et al., Dermonecrosis caused by a spitting cobra snakebite results from toxin potentiation and is prevented by the repurposed drug varespladib. Proc Natl Acad Sci U S A, 2024. 121(19): p. e2315597121.

9. Tasoulis, T. and G.K. Isbister, A current perspective on snake venom composition and constituent protein families, in Archives of Toxicology. 2023, Springer Science and Business Media Deutschland GmbH. p. 133–153.

10. Mackessy, S.P., Venom Ontogeny in the Pacific Rattlesnakes Crotalus Viridis Helleri and C. v. Oreganus. Copeia, 1988. 1988(1): p. 92–101.

11. Lomonte, B., et al., Neutralizing interaction between heparins and myotoxin II, a lysine 49 phospholipase A2 from Bothrops asper snake venom. Identification of a heparin-binding and cytolytic toxin region by the use of synthetic peptides and molecular modeling. Journal of Biological Chemistry, 1994. 269(47): p. 29867–29873.

12. Kini, R.M. and H.J. Evans, A model to explain the pharmacological effects of snake venom phospholipases A2. Toxicon, 1989. 27(6): p. 613–35.

13. Lewin, M.R., et al., Varespladib (LY315920) Appears to Be a Potent, Broad-Spectrum, Inhibitor of Snake Venom Phospholipase A2 and a Possible Pre-Referral Treatment for Envenomation. Toxins, 2016. 8(9): p. 248–248.

14. Lewin, M.R., et al., Varespladib in the Treatment of Snakebite Envenoming: Development History and Preclinical Evidence Supporting Advancement to Clinical Trials in Patients Bitten by Venomous Snakes. Toxins (Basel), 2022. 14(11).

15. Lewin, M.R., et al., Delayed LY333013 (Oral) and LY315920 (Intravenous) Reverse Severe Neurotoxicity and Rescue Juvenile Pigs from Lethal Doses of Micrurus fulvius (Eastern Coral Snake) Venom. Toxins, 2018b. 10(11).

16. Tan, C.H., T.M.C. Lingam, and K.Y. Tan, Varespladib (LY315920) rescued mice from fatal neurotoxicity caused by venoms of five major Asiatic kraits (Bungarus spp.) in an experimental envenoming and rescue model. Acta Tropica, 2022. 227(September 2021): p. 106289–106289.

17. Dawson, C.A., et al., A drug combination therapy consisting of toxin phospholipase A2 and metalloproteinase inhibitors provides preclinical protection against North American Crotalid snakebite envenoming. bioRxiv, 2025.

18. Rudresha, G.V., et al., Preclinical evaluation of small molecule inhibitors as early intervention therapeutics against Russell’s viper envenoming in India. Communications Medicine, 2025. 5(1): p. 226.

19. Gerardo, C.J., et al., Oral varespladib for the treatment of snakebite envenoming in India and the USA (BRAVO): a phase II randomised clinical trial. BMJ Global Health, 2024. 9(10).

20. Dart, R.C., et al., Validation of a severity score for the assessment of crotalid snakebite. Annals of Emergency Medicine, 1996. 27(3): p. 321–326.

21. Xie, C., et al., Varespladib Inhibits the Phospholipase A2 and Coagulopathic Activities of Venom Components from Hemotoxic Snakes. Biomedicines, 2020b. 8(6): p. 165–165.

22. Albulescu, L.-O., et al., A therapeutic combination of two small molecule toxin inhibitors provides broad preclinical efficacy against viper snakebite. Nature Communications, 2020. 11(1): p. 6094–6094.

23. Vergara, I., et al., Eastern Coral Snake Micrurus fulvius Venom Toxicity in Mice is Mainly Determined by Neurotoxic Phospholipases A2. Journal of Proteomics, 2014. 105: p. 295–306.

24. Gutierrez, J.M. and C.L. Ownby, Skeletal muscle degeneration induced by venom phospholipases A2: insights into the mechanisms of local and systemic myotoxicity. Toxicon, 2003. 42(8): p. 915–31.

25. Bhalla, A., et al., An Experience with Manual Ventilation in Respiratory Paralysis Due to Indian Common Krait (Bungarus caeruleus) Bite. Asia Pacific Journal of Medical Toxicology, 2014. 3(2): p. 55–58.

26. Talukdar, A. and R. Doley, Identification of poorly immunodepleted phospholipase A2 (PLA2) proteins of Bungarus fasciatus venom from Assam, India and evaluation of Indian polyvalent antivenom using third-generation antivenomics. Toxicon, 2024. 239.

27. Gerardo, C.J., et al., Does This Patient Have a Severe Snake Envenomation?: The Rational Clinical Examination Systematic Review, in JAMA Surgery. 2019, American Medical Association. p. 346-354.

