## Supplemental Data for "Intravenous Followed by Oral Varespladib as Late Adjunctive Therapy for Snakebite: a Phase II Randomized Clinical Trial"

### Supplemental Appendix

**Appendix 1.** List of sites in India and the U.S. for the BRAVIO trial.

India

1. S. P. Medical College, Bikaner, Rajasthan
2. Post Graduate Institute of Medical Education & Research (PGIMER), Chandigarh
3. Jawaharlal Institute of Postgraduate Medical Education & Research (JIPMER), Puducherry
4. K. R. Hospital, Mysore Medical College & Research Insititute, Mysuru, Karnataka
5. All India Institute of Medical Sciences, Raipur, Chhattisgarh
6. King George University Hospital, Lucknow, Uttar Pradesh
7. Nil Ratan Sircar Medical College, Kolkata, West Bengal
8. All India Institute of Medical Sciences, Bhubaneshwar, Odisha

United States

9. Duke University, Durham, North Carolina
10. University of Kentucky, Lexington, Kentucky
11. University of Arizona, Tucson, Arizona
12. Banner - University Medical Center, Phoenix, Arizona
13. Tampa General Hospital, Tampa, Florida
14. Antelope Valley Medical Center, Lancaster, California
15. Texas Tech University Health Sciences, El Paso, Texas
16. Desert Regional Medical Center, Palm Springs, California
17. University of Texas Health-San Antonio, San Antonio, Texas
18. University of Florida Health, Gainesville, Florida

**Appendix 2.** Snakebite Severity Score versions used for (i) Inclusion Criteria; (ii) Primary Outcome (Viper); (iii) Secondary Outcomes (SSS AUC basline to Day 7 and complete recovery).

| | |
|---|---|
| **Local wound** [a b c] | |
| No signs/symptoms | 0 |
| Pain, swelling, or ecchymosis within 5–7.5 cm of bite site | 1 |
| Pain, swelling, or ecchymosis involving less than half the extremity (7.5–50 cm from bite site) | 2 |
| Pain, swelling, or ecchymosis involving half to all of extremity (50–100 cm from bite site) | 3 |
| Pain, swelling, or ecchymosis extending beyond affected extremity (more than 100 cm of bite site) | 4 |
| **Pulmonary system** [a c] | |
| No signs/symptoms | 0 |
| Dyspnea, minimal chest tightness, mild/vague discomfort, respirations of 20–25 breaths per minute | 1 |
| Moderate respiratory distress, 26–40 bpm | 2 |
| Cyanosis, air hunger, extreme tachypnea, or respiratory insufficiency /failure | 3 |
| **Cardiovascular system** [a c] | |
| No signs/symptoms | 0 |
| HR 100–125 BPM, palpitations, generalized weakness, benign dysrhythmia, or hypertension | 1 |
| HR 126–175 BPM, or hypotension with SBP > 100 mmHg | 2 |
| HR > 175 BPM, or hypotension with SBP < l00 mmHg, malignant dysrhythmia, or cardiac arrest | 3 |
| **Gastrointestinal system** | |
| No signs/symptoms | 0 |
| Pain, tenesmus, or nausea | 1 |
| Vomiting or diarrhea | 2 |
| Repeated vomiting, diarrhea, hematemesis, or hematochezia | 3 |
| **Hematologic system** [a b c] | |
| No signs/symptoms | 0 |
| Coagulation parameters slightly abnormal: PT ULN–20 secs, PTT ULN–50 secs, platelets 100–150K/mL, or fibrinogen 100–150 mcg/mL | 1 |
| Coagulation parameters abnormal: PT 20–50 secs, PTT 50–75 secs, platelets 50–100K/mL, or fibrinogen 50–100 mcg/mL | 2 |
| Coagulation parameters abnormal: PT 50–100 secs, PTT 75–100 secs, platelets 20–50K/mL, or fibrinogen < 50 mcg/mL | 3 |
| Coagulation parameters markedly abnormal, with serious bleeding or the threat of spontaneous bleeding; unmeasurable PT or PTT, platelets < 20 K/mL, undetectable fibrinogen, severe abnormalities of other laboratory values also fall into this category | 4 |
| **Nervous system** [a b c] | |
| No signs/symptoms | 0 |
| Minimal apprehension, headache, weakness, dizziness, chills, or paresthesia | 1 |
| Moderate apprehension, headache, weakness, dizziness, chills, paresthesia, confusion, fasciculation in area of bite site, ptosis, or dysphagia | 2 |
| Severe confusion, lethargy, weakness, paralysis, seizures, coma, psychosis, or generalized fasciculation | 3 |
| **Renal system** [c] | |
| Normal creatinine and urine output | 0 |
| Creatinine 1.5 to 1.9 times baseline, increase in creatinine ≥0.3mg/dl (≥26.5 μmol/L) from baseline, or urine output <0.5 ml/kg/h for >6 h | 1 |
| Creatinine 2 to 2.9 times baseline or urine output <0.5 ml/kg/h for >12 h | 2 |
| Creatinine ≥3.0 times baseline, increase in creatinine to ≥4.0 mg/dl (≥353.6 μmol/L), urine output <0.3 ml/kg/h for ≥24 h or anuria ≥12 h, or initiation of renal replacement therapy | 3 |
| **TOTAL** | |

[a] Subscores used for inclusion determination. Hematologic subscore may be used if available, but enrollment should not be delayed. Abnormal 20WBCT may be used for inclusion.
[b] Subscores used for primary endpoint for viper bites (3-item SSS AUC baseline to Day 14). Definitive laboratory testing should be used for efficacy endpoints.
[c] Subscores used for key secondary outcomes (6-item SSS AUC baseline to Day 7 and complete recovery). Definitive laboratory testing should be used for efficacy endpoints.

**Appendix 3:** Complete eligibility criteria.

<u>Inclusion Criteria</u>

1. Is a male or female ≥ 5 years of age with venomous snakebite.
2. Patients must have known or suspected venomous snakebite. In India, enrollment will be restricted to patients bitten by suspected or confirmed Russell's viper (*Daboia russelii*) or krait (*Bungarus* spp.). In the U.S., any snakebite that meets all other criteria may be eligible.
3. Patients must meet one of two categories of inclusion criteria:
   - Category 1: The patient is enrolled within 5 hours of venomous snake bite or symptom onset with an SSS score* of ≥2 in one system and ≥1 in another system (2+1). OR
   - Category 2: The patient has a suspected or confirmed bite from an elapid and is enrolled within 10 hours of bite or symptom onset with moderate to severe cranial nerve or skeletal muscle weakness**.
4. Is willing (or legally authorized representative is willing) to provide informed consent prior to initiation of any study procedures.

*Only local wound, pulmonary, cardiovascular, haematologic, or nervous system scores qualify for SSS inclusion criteria. GI and Renal scores are not used for inclusion. Haematologic score may be counted if available, but inclusion should not wait for laboratory results. Point of care tests (e.g., 20WBCT) may be used for enrollment, if used per site standard of care. GI and Renal scores are not used for inclusion.
** Isolated ptosis does not meet the definition of moderate to severe neurotoxicity for enrollment

<u>Exclusion Criteria</u>

1. Has history of or is suspected to have cerebrovascular accident or intracranial bleeding of any kind, acute coronary syndrome, myocardial infarction, or severe pulmonary hypertension.
2. Has known history of inherited bleeding or coagulation disorder.
3. Is, at Screening Visit, using the following anticoagulants: warfarin/coumadin, argatroban, bilvalirudin, lepirudin, apixaban, dabigatran, clopidogrel, prasugrel, ticlodipine or another anticoagulant agent not specifically listed, or has used heparin, enoxaparin, fondaparinux, or other low molecular weight heparin or any antiarrhythmic drugs within 14 days prior to treatment.
4. Has a history of chronic liver disease such as chronic active viral hepatitis, alcohol-related liver disease, non-alcoholic steatohepatitis, non-alcoholic fatty liver disease, hemochromatosis, primary biliary cirrhosis, primary sclerosing cholangitis, autoimmune hepatitis.
5. Reports or has known pre-existing renal impairment or chronic kidney disease.
6. Has a known allergy or significant adverse reaction to varespladib or varespladib-methyl.
7. Is considered by the Investigator to be unable to comply with protocol requirements due to geographic considerations, psychiatric disorders, or other compliance concerns.
8. Is pregnant, has a positive urine or serum human chorionic gonadotropin (hCG) pregnancy test or not willing to use a highly effective method of contraception for 14 days after initial treatment, or is breast-feeding.

**Appendix 4:** Details of the sample size calculation for patients bitten by elapids.

For patients bitten by elapids, calculation of power was based on a randomisation test. Because the time to head lift of 5 seconds is measured at discrete time intervals, when a person first lifts his or her head for 5 seconds, the assigned time will be assumed to be half-way between the previous time of assessment and the time of successful head lift. The expected percentage of placebo and varespladib subjects who first lift their heads at specific times, as well as the assigned times, are shown below. The percentages for placebo and for varespladib are based on the literature, experience from OPX-PR-01 (BRAVO Trial), and experience of the investigators.

| | | **Placebo** | | | **Varespladib** | |
|---|---|---|---|---|---|---|
| **Observed time of first 5 sec head lift** | **Assigned time (hours)** | **%** | **Assigned Time × %** | | **%** | **Assigned Time × %** |
| **3 hours** | 1.5 | 0 | 0 | | 8 | 12 |
| **6 hours** | 4.5 | 2 | 9 | | 12 | 54 |
| **24 hours** | 15 | 38 | 570 | | 60 | 900 |
| **Day 3** | 36 | 45 | 1620 | | 20 | 720 |
| **Day 7** | 108 | 13 | 1404 | | 0 | 0 |
| **Day 14** | 252 | 2 | 504 | | 0 | 0 |
| **Day 28** | 504 | 0 | 0 | | 0 | 0 |
| **Mean time** | | | 41.07 | | | 16.86 |

To calculate power, we assumed the study will have 40 subjects with elapid bites. We simulated the above placebo distribution 200 times for 40 subjects then randomly split the assignments so that 20 each would receive placebo and varespladib. Roughly 95% of these simulated distributions found a difference between the two groups of less than 24.21 (that is, 41.07 – 16.86). We therefore conclude that under the above assumptions for the distribution of results, the power of the test of the elapids is above 90%.

**Appendix 5:** Multiple imputation methodology.

The multiple imputation (MI) procedures for the primary and key secondary endpoints are described below. For the primary analysis, missing data not due to mortality will be imputed using the following rules. No imputation will be performed to define the time to complete recovery for the 5-second head-lift. We anticipate all patients will have these data.

Relevant to patients bitten by vipers, the following imputation algorithm will be applied to each SSS subscore separately (hemotoxicity, local wound, neurotoxicity). The primary outcome SSS score will be the sum of the observed and imputed scores at each assessment time point.

1. Intermittent missing values (i.e. missing data prior to the first intercurrent event or any missing data in patients without an intercurrent event) will be imputed under the missing at random (MAR) assumption within each treatment group using the Markov Chain Monte Carlo (MCMC) Methodology to create partially imputed datasets which have a monotone missing structure. The parameters to be included in the imputation model are Strata (study site, snake type) and SSS subscore values at each assessment timepoint from baseline (Day 1, Pre-dosing) through to Day 14.
2. Having achieved a monotone pattern, scores at each analysis visit from baseline to Day 14 will be imputed sequentially in the order specified in the VAR statement of the MI procedure. For this, the Predictive Mean Matching method (PMM) will be used. This method is particularly helpful if the normality assumption is violated. For subjects with complete data up to a particular visit, a PMM model will be fit that includes the outcome at that visit as the dependent variable. The independent variables will be treatment assignment, study site, snake type, and non-missing scores. The seed will be 931428. This process will be repeated 15 times, resulting in a total of 150 complete analysis datasets.
3. For each completed dataset, any necessary derived variables will be computed. Then the ANCOVA model will be performed for the SSS AUC from baseline to Day 14. The results will be combined into one MI inference (LS mean, associated 95% CI, and p-value) using PROC MIANALZE as illustrated by Ratitch et al., 2013.

The first key secondary analysis (SSS AUC Day 7) will use the same process above, except in step 1 the scores for cardiovascular toxicity, pulmonary toxicity, and renal toxicity will also be evaluated, and in step 3 the AUC based on the 6 subscores from baseline to Day 7 will be derived for each subject in each imputed dataset, then analyzed and aggregated as above using PROC MIANALZE. (Mallinckrodt et al., 2013)

The second key secondary analysis (complete SSS recovery at Day 28) will use the same process as describe above, except in step 3 the analysis will use logistic regression. Non-integer scores between 0 and 1 at Day 28 will be rounded to 0 or 1 using “round half up” rounding rules (i.e., 0.5 will be rounded to 1 and categorized as not completely recovered, values less than 0.5 will be categorized as completely recovered).

The third key secondary analysis (PSFS at Day 3) has the potential for missing data at baseline (a covariate in the analyses) and missing data at Day 3 (the outcome). If there are missing data at baseline, the earliest reported PSFS score in the first 24 hours will be considered as representative of the baseline score. Patients who have missing PSFS data at all time points from baseline through 24 hours will not be included in the primary analysis. A sensitivity analysis will be conducted in which the overall sample average baseline PSFS is used for patients with missing baseline through 24-hour PSFS data. Missing data at Day 3 will be imputed using the same MI approach as the primary endpoint. The independent variables will be treatment assignment, study site, snake type, and non-missing PSFS scores. Because treatment is a key variable in the imputation model, we will not impute Day 3 PSFS for patients who do not complete the first 6 hours of infusion of the study drug.

Ratitch, B., O'Kelly, M., & Tosiello, R. (2013). Missing data in clinical trials: From clinical assumptions to statistical analysis using pattern mixture models. *Pharmaceutical Statistics*, *12*(6). https://doi.org/10.1002/pst.1549

**Appendix 6:** Key secondary outcomes in pre-specified subgroups.

| Outcome | Varespladib | Placebo | Treatment Effect (95% CI) |
|---|---|---|---|
| **Patients in India** | **44** | **42** | |
| 6-item SSS AUC Day7, Mean (SE) | 406.3 (32.6) | 434.1 (32.9) | -27.8 (-118.6 to 63.0) |
| Complete recovery, % | 57% | 58% | 1% (-21% to 21%) |
| Patient Specific Functional Scale Day 3, Mean (SE) | 7.3 (0.5) | 6.9 (0.5) | 0.4 (-0.9 to 1.6) |
| **Patients in U.S.** | **29** | **24** | |
| 6-item SSS AUC Day7, Mean (SE) | 429.3 (39.3) | 496.4 (44.2) | -67.1 (-183.1 to 48.8) |
| Complete recovery, % | 54% | 48% | -6% (-22% to 32%) |
| Patient Specific Functional Scale Day 3, Mean (SE) | 5.5 (0.6) | 5.0 (0.7) | 0.5 (-1.3 to 2.3) |
| **Patients randomized ≤5hrs after bite**[a] | **38** | **28** | |
| 6-item SSS AUC Day7, Mean (SE) | 467.0 (38.9) | 464.9 (43.9) | 2.1 (-102.7 to 106.9) |
| Complete recovery, % | 56% | 50% | 6% (-19% to 30%) |
| Patient Specific Functional Scale Day 3, Mean (SE) | 6.05 (0.6) | 6.03 (0.6) | 0.02 (-1.5 to 1.6) |
| **Patients randomized >5hrs after bite** | **35** | **38** | |
| 6-item SSS AUC Day7, Mean (SE) | 407.0 (49.4) | 487.2 (45.4) | -80.2 (-173.3 to 12.8) |
| Complete recovery, % | 55%<br>19/35 | 53%<br>20/38 | 2% (-21% to 25%) |
| Patient Specific Functional Scale Day 3, Mean (SE) | 6.1 (0.7) | 5.2 (0.7) | 0.9 (-0.5 to 2.3) |

**Appendix 7.** Adverse events and selected laboratory abnormalities. Values represent the number (%) of patients with one or more adverse events or laboratory abnormalities.

| **Safety Outcome - no. (%)** | **Varespladib (n=73)** | **Placebo (n=66)** |
|---|---|---|
| Treatment-Emergent Adverse Events | 19 (26) | 20 (30) |
| Related Treatment-Emergent Adverse Events | 3 (4) | 7 (11) |
| Serious Adverse Events | 0 (0) | 2 (3) |
| Adverse Events by System Organ Class | | |
| Gastrointestinal disorders | 4 (6) | 2 (3) |
| Infections | 2 (3) | 4 (6) |
| Acute kidney injury | 0 | 1 (2) |
| Hepatic enzyme increase | 0 | 2 (3) |
| Cardiac disorders | 0 | 1 (2) |
| Skin/subcutaneous tissue disorders | 1 (1) | 2 (3) |
| Nervous System Disorders | 7 (10) | 5 (8) |
| Laboratory Abnormalities | | |
| Elevated creatine* | 7 (10) | 6 (9) |
| Elevated liver enzymes** | 2 (3) | 8 (12) |

* Defined as a creatinine at any timepoint post-baseline either ≥ 0.3 mg/dL or ≥ 1.5 times baseline creatinine.

** Defined as post-baseline liver enzymes, either AST or ALT, exceeding 3x the upper limit of normal.
